# Projecting the impact of a two-dose COVID-19 vaccination campaign in Ontario, Canada

**DOI:** 10.1101/2020.12.10.20246827

**Authors:** Thomas N. Vilches, Kevin Zhang, Robert Van Exan, Joanne M. Langley, Seyed M. Moghadas

## Abstract

**Background:** A number of highly effective COVID-19 vaccines have been developed and approved for mass vaccination. We evaluated the impact of vaccination on COVID-19 outbreak and disease outcomes in Ontario, Canada.

**Methods:** We used an agent-based transmission model and parameterized it with COVID-19 characteristics, demographics of Ontario, and age-specific clinical outcomes. We implemented a two-dose vaccination program according to tested schedules in clinical trials for Pfizer-BioNTech and Moderna vaccines, prioritizing healthcare workers, individuals with comorbidities, and those aged 65 and older. Daily vaccination rate was parameterized based on vaccine administration data. Using estimates of vaccine efficacy, we projected the impact of vaccination on the overall attack rate, hospitalizations, and deaths. We further investigated the effect of increased daily contacts at different stages during vaccination campaigns on outbreak control.

**Results:** Maintaining non-pharmaceutical interventions (NPIs) with an average of 74% reduction in daily contacts, vaccination with Pfizer-BioNTech and Moderna vaccines was projected to reduce hospitalizations by 27.3% (95% CrI: 22.3% - 32.4%) and 27.0% (95% CrI: 21.9% - 32.6%), respectively, over a one-year time horizon. The largest benefits of vaccination were observed in preventing deaths with reductions of 31.5% (95% CrI: 22.5% - 39.7%) and 31.9% (95% CrI: 22.0% - 41.4%) for Pfizer-BioNTech and Moderna vaccines, respectively, compared to no vaccination. We found that an increase of only 10% in daily contacts at the end of lockdown, when vaccination coverage with only one dose was 6%, would trigger a surge in the outbreak. Early relaxation of population-wide measures could lead to a substantial increase in the number of infections, potentially reaching levels observed during the peak of the second wave in Ontario.

**Conclusions:** Vaccination can substantially mitigate ongoing COVID-19 outbreaks. Sustaining population-wide NPIs, to allow for a sufficient increase in population-level immunity through vaccination, is essential to prevent future outbreaks.

## Introduction

Despite unprecedented public health measures, such as stay-at-home orders, school closures, and physical distancing (1–4), the novel coronavirus disease 2019 (COVID-19) has caused severe global health and tremendous economic consequences (5,6). In Canada, these measures have been comparatively effective in flattening initial outbreaks (7,8). However, despite continued non-pharmaceutical interventions (NPIs), the second wave proved to be a deadly outbreak, underscoring the importance of a safe and effective vaccine to control the ongoing pandemic.

With an accelerated pace of vaccine development, several vaccines have received approval after demonstrating relatively high efficacy in clinical trials (9). In Canada, Pfizer-BioNTech and Moderna vaccines were authorized for distribution in December 2020 (10). However, shortages in early vaccine supply hindered efforts to suppress the second wave, triggering lockdown in Canadian provinces. With improving rates of vaccination and increasing supply due to additional authorization of vaccines (i.e., Oxford-AstraZeneca and Janssen), there is a need to understand the potential impact of vaccination campaigns with vaccine prioritization (11), and timelines for lifting restrictive NPIs.

We sought to evaluate the impact of a COVID-19 vaccination campaign, based on a scenario with two doses of Pfizer-BioNTech or Moderna vaccines distributed, 21 or 28 days apart, on the overall attack rate, hospitalizations, and deaths in Ontario, the most populated Canadian province. We extended an agent-based model of disease transmission (12) to include vaccination with an age-specific uptake distribution prioritized based on recommendations outlined by the National Advisory Committee on Immunization (13,14). We evaluated a roll-out strategy that prioritizes high risk adults (i.e., healthcare workers, elderly, and comorbid individuals), followed by the general population, to reduce transmission and severe outcomes (11,13).

## Methods

### Model structure

We extended a previously established agent-based COVID-19 transmission model (12) and included vaccination to simulate outbreak scenarios. The natural history of COVID-19 was implemented in the model by considering individual status as susceptible; latently infected (not yet infectious); asymptomatic (infected and infectious but with no symptoms); pre-symptomatic (infected, infectious and in the stage before symptomatic illness); symptomatic with either mild or severe/critical illness; recovered (and not infectious); and dead (Appendix, Figure A1). We binned the model population into six age groups of 0-4, 5-19, 20-49, 50-64, and 65-79, and 80+ years old based on the demographics of Ontario, Canada (15) and parameterized the model with estimates of the proportion of the population with comorbidities associated with severe COVID-19 (Appendix, Table A1) to allow for the implementation of vaccine prioritizations (16,17). We assumed the same degree of susceptibility to infection across all age groups. Interactions between individuals were informed using an empirically determined contact network (18). The daily number of contacts for each individual was sampled from a negative-binomial distribution with age-dependent mean and standard deviation (Appendix, Table A2). The daily number of contacts during lockdown or self-isolation was reduced by an average of 74%, based on a matrix derived from a representative sample population during COVID-19 lockdown (19).

### Disease dynamics

We implemented disease transmission in a probabilistic manner whereby susceptible individuals were exposed to infectious individuals (i.e., asymptomatic, pre-symptomatic, or symptomatic stages of the disease). Infected individuals entered a latent period as part of an average incubation period of 5.2 days (20). For those who went on to develop symptomatic disease, the incubation period included a pre-symptomatic stage prior to the onset of symptoms (21), with a mean duration of 2.3 days sampled from a Gamma distribution with a mean (21). The infectious period post-symptom onset was also sampled from a Gamma distribution with a mean of 3.2 days (22). We considered an age-dependent probability of developing mild, severe, or critical illness after symptom onset. Infected individuals who did not develop symptoms remained asymptomatic after the latent period until recovery. Asymptomatic individuals were infectious for an average of 5 days, which was sampled from a Gamma distribution (22,23).

Based on the number of secondary cases generated during each stage of the disease (24), we parameterized the infectivity of asymptomatic, mild symptomatic, and severe symptomatic stages to be 26%, 44%, and 89% relative to the pre-symptomatic stage (24–26). We assumed that recovered individuals could not be re-infected during the same outbreak scenario.

### Infection outcomes

In our model, mild symptomatic cases recovered without the need for hospitalization. A proportion of individuals with severe illness used hospital beds in this model. We parameterized the model for the use of intensive care unit (ICU) and non-ICU beds based on recent COVID-19 hospitalization data stratified by the presence of comorbidities in Ontario (17). We assumed that all symptomatic cases who were not hospitalized self-isolated within 24 hours of symptom onset for the entire symptomatic period. The time from symptom onset to hospital admission was uniformly sampled in the range of 2 to 5 days (12,27). The lengths of non-ICU and ICU stays were sampled from Gamma distributions with means of 12.4 and 14.4 days, respectively (28,29).

### Vaccination

We implemented a two-dose vaccination campaign, with a daily vaccination rate that increased over time and saturated at 50 doses per day per 10,000 population (Appendix, Figure A2).

Vaccination was sequential with prioritization of: (i) healthcare workers, individuals with comorbidities, and those aged 65 and older (i.e., protection cohort); followed by (ii) individuals aged 16-64 (i.e., disruption minimization cohort) (11,13). Children under 16 years of age were not vaccinated. In our model, the age-eligibility was 16+ for Pfizer-BioNTech vaccines, but 18+ for Moderna. We assumed that the maximum achievable coverage within a one-year time horizon was 95% among healthcare workers and those aged 65 and older. The maximum coverage was set to 70% among other age groups (Appendix, Table A3). Pre-existing immunity as a result of a primary infection with COVID-19 was not a factor in the vaccination of individuals.

For Pfizer-BioNTech vaccines, we assumed vaccination campaigns followed the recommended dosing interval (i.e., tested in clinical trials) of 21 days (30). This interval was 28 days for Moderna vaccines (31). Vaccine coverage of the entire population with two doses reached 62% for both Moderna and Pfizer-BioNTech within one year of the vaccination campaign.

We reviewed published studies and briefing documents for vaccine approvals on the efficacy of both Pfizer-BioNTech and Moderna vaccines in preventing infection, symptomatic disease, and severe disease (30–35). We implemented the estimated efficacies and associated timelines following the first dose of vaccine for each vaccinated individual (Appendix, Table A4).

### Model scenarios and implementation

We considered a 5% level of pre-existing immunity accrued prior to the start of simulations (corresponding to October 1, 2020) based on seroprevalence studies (8,36). To distribute this level of population immunity, we first simulated the model in the absence of vaccination and derived the infection rates in different age groups when the overall attack rate reached 5% of the population. We then used the age-specific attack rates to initialize the proportion of the population with immunity in different age groups for the vaccination model.

We calibrated the transmission probability per contact to the effective reproduction number R_e_=1.12 estimated in early October 2020, accounting for the effect of NPIs in Ontario (37). With this calibration and using parameters provided in Table 1 and Table A5 (Appendix), we fitted the model to the reported cases per 10,000 population over time from October 1, 2020 to March 8, 2021. Vaccination started on day 74 of the simulations (i.e., December 14, 2020). We averaged the results for the overall attack rate, hospitalizations, and deaths over 1000 independent

**Table 1.**
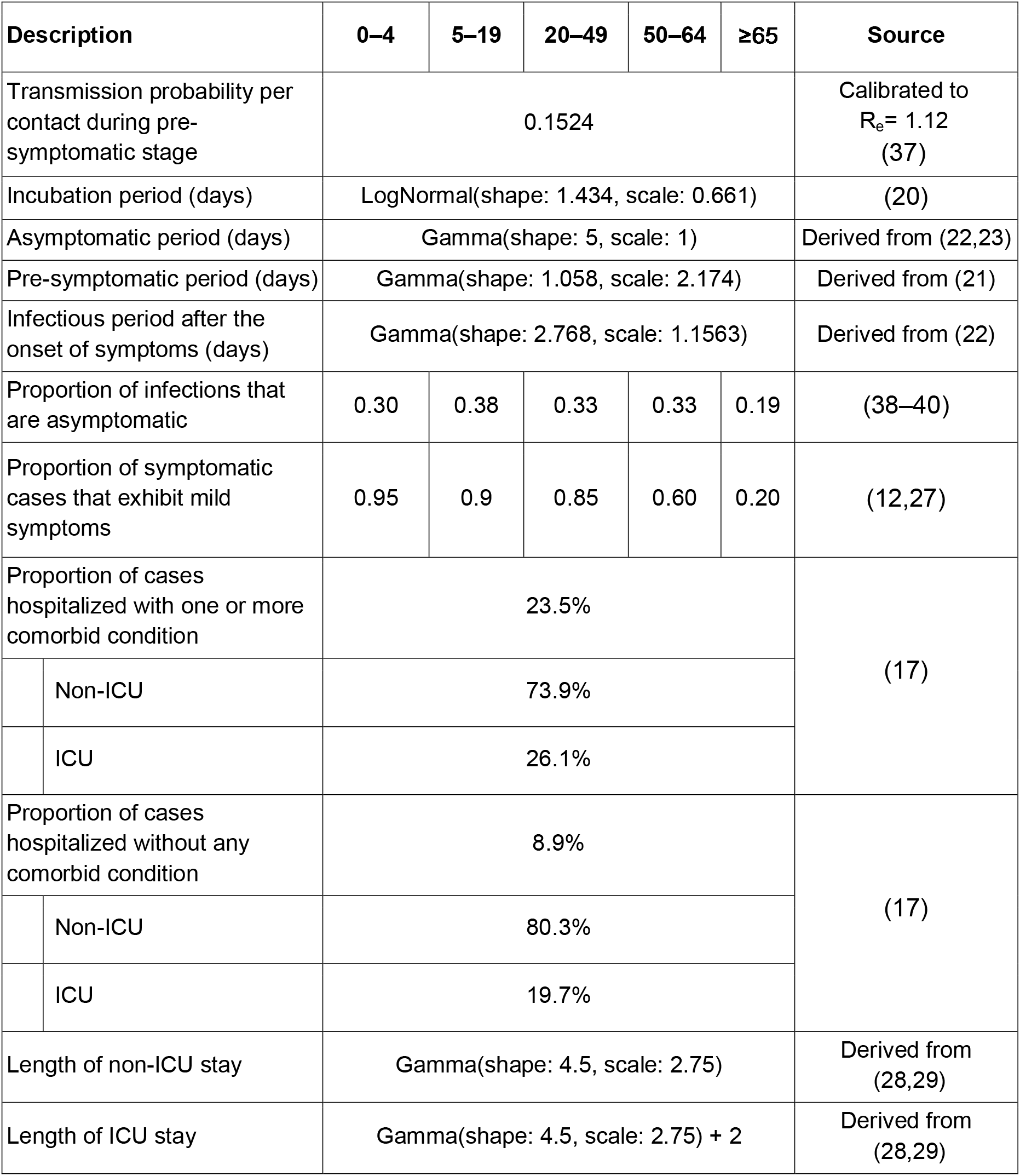
Description of model parameters and their estimates.

Monte-Carlo realizations, and derived the credible intervals (at the 5% significance level) for reduction of attack rate and severe outcomes attributable to vaccination using a bias-corrected and accelerated bootstrap method (with 500 replications). The model was coded in Julia language and is available at: https://github.com/thomasvilches/vac_covid_ontario.

## Results

From the start of vaccination on day 74 of simulations, we projected an overall attack rate of 1.7% (95% CrI: 1.6% - 1.9%) using Pfizer-BioNTech or Moderna vaccines, if the daily number of contacts did not change after the lockdown (stay-at-home order) period ended on day 158 (March 8, 2021) (Figure 1A). Maintaining NPIs in place, the reduction of attack rate attributable to vaccination was projected to be 26.4% (95% CrI: 22.0% - 30.7%) for Pfizer-BioNTech and 27.5% (95% CrI: 22.6% - 31.6%) for Moderna vaccines over a one-year time horizon (Figure 1B). The corresponding reductions of hospitalizations were 27.3% (95% CrI: 22.3% - 32.4%) and 27.0% (95% CrI: 21.9% - 32.6%). The largest benefits of vaccination were observed in preventing deaths with reductions of 31.5% (95% CrI: 22.5% - 39.7%) and 31.9% (95% CrI: 22.0% - 41.4%) for Pfizer-BioNTech and Moderna vaccines, respectively, compared to no vaccination (Figure 1B).

**Figure 1.**
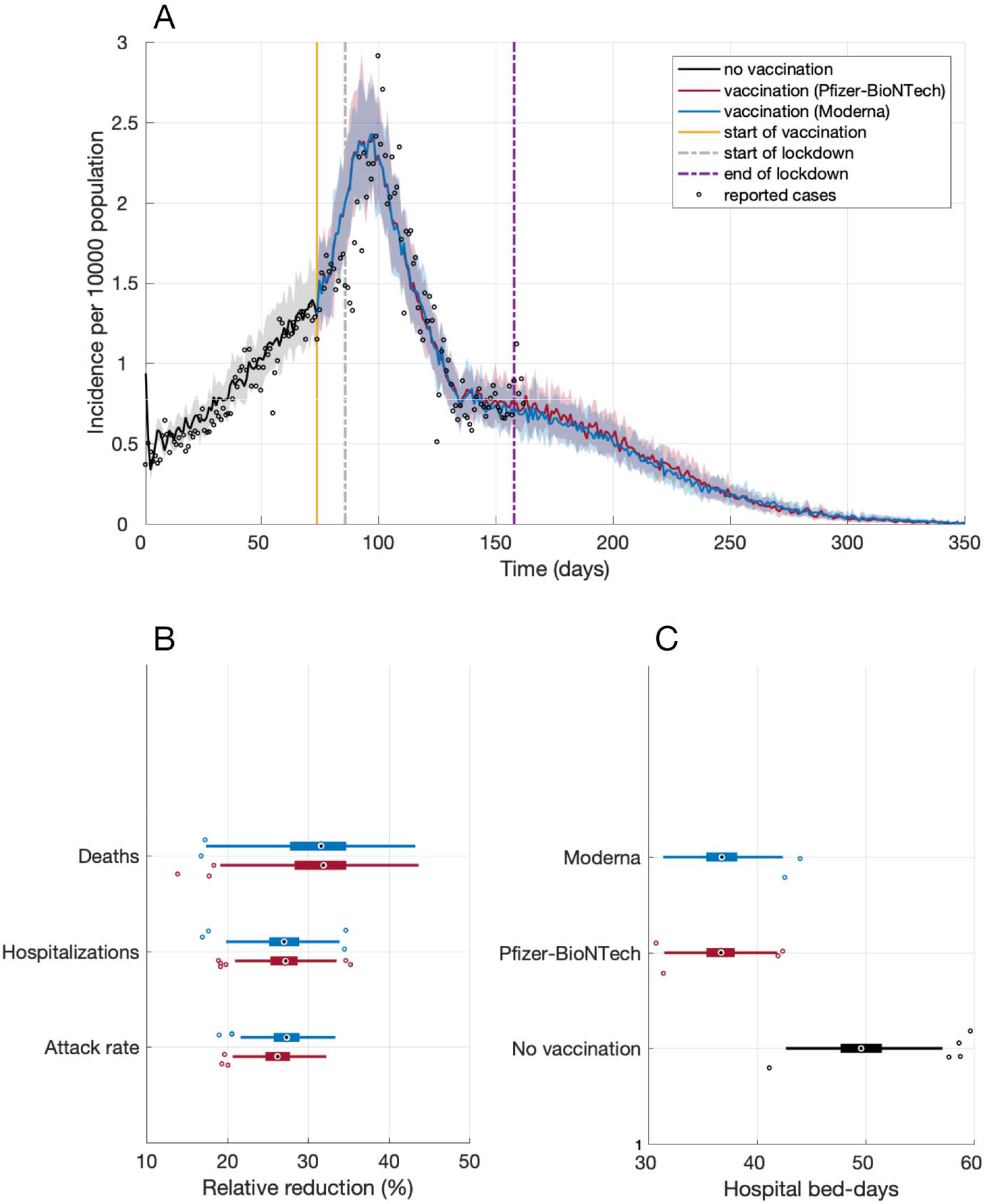
(A) Projected incidence of infection with fitting to reported cases per 10,000 population from October 1, 2020 (day 0) to March 12, 2021 (day 162). (B) Projected reduction of overall attack rate, hospitalizations and deaths from the start of vaccination (day 74) compared to no vaccination for a time horizon of one year. (C) Projected number of hospital bed-days per 10,000 population from day 74 for one year. Box plots indicate interquartile range (IQR), and horizontal lines are the extended range from minimum (25th percentile – 1.5 IQR) to maximum (75th percentile + 1.5 IQR).

If vaccines were not available, we projected an average of 49.9 (95% CrI: 44.3 - 55.6) hospital bed-days per 10,000 population from day 74 onwards (Figure 1C). With vaccination, hospital bed-days reduced significantly to 36.7 (95% CrI: 32.6 - 40.9) and 36.7 (95% CrI: 32.8 - 40.6) per 10,000 population for Pfizer-BioNTech and Moderna vaccines, respectively.

### Effect of vaccination with increased daily contacts

We simulated the effect of vaccination with increased daily number of contacts following the lockdown period, accounting for possible reduction in the adherence to NPIs. At the end of lockdown (day 158 of simulations), vaccination coverage with only one dose was 6% for both Pfizer-BioNTech and Moderna vaccines. At the same time, vaccination coverage of those who had received both doses was 1.7% and 1.9% for Pfizer-BioNTech and Moderna vaccines, respectively. If the daily number of contacts was increased by 10% immediately after lockdown ended (March 8, 2021), we observed a gradual increase in the case incidence, reaching a peak of 0.93 cases (on average) per 10,000 population within 40 days (Figure 2A). Increasing the daily number of contacts by 20% would lead to a significant increase in the daily incidence of infection, reaching a peak of 1.82 (on average) per 10,000 population within 60 days similar to the increasing incidence trend observed in late December 2020 during the second wave, which triggered the instigation of province-wide lockdown (Figure 2D).

**Figure 2.**
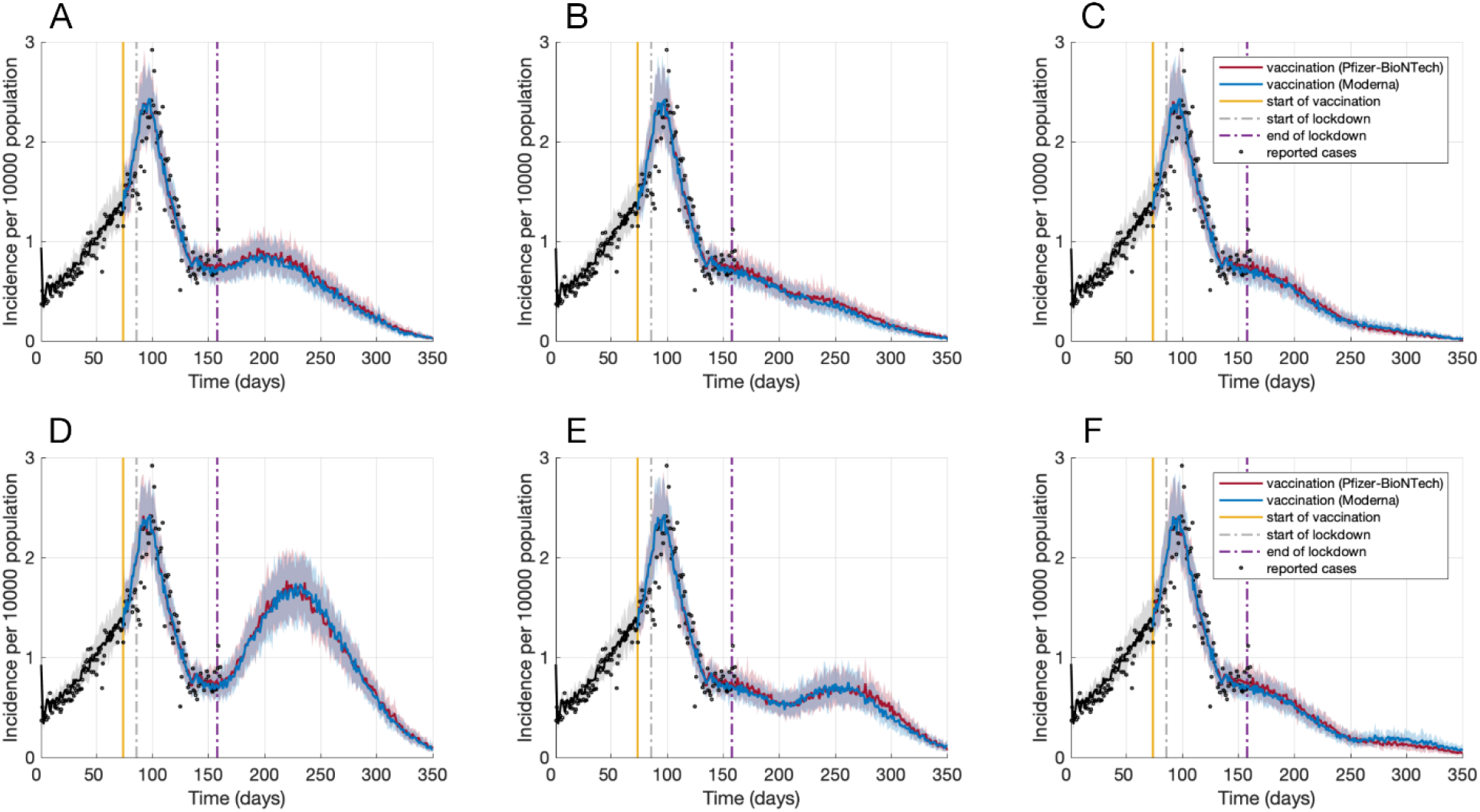
Projected incidence of infection per 10,000 population post-lockdown with a 10% increase in the daily number of contacts on day 158 (A), 200 (B), or 250 (C), and with a 20% increase in the daily number of contacts on day 158 (D), 200 (E), or 250 (F).

If contacts post-lockdown did not change until day 200 (April 19, 2021), then a 10% increase in daily number of contacts had no significant impact on the declining trend of the outbreak (Figure 2B). On day 200, vaccination coverage with only one dose was 23%, with coverage of 8.4% and 8.0% for both doses of Pfizer-BioNTech and Moderna vaccines, respectively. However, these vaccine coverages are still insufficient to prevent a surge in infections associated with larger increases in the daily number of contacts. For example, a 20% increase in contacts would lead to an increasing trajectory of the outbreak (Figure 2E), approaching a peak with an incidence similar to that reported at the end of the lockdown period.

If contacts did not change until day 250 (June 10, 2021), there was no surge in the number of cases with a 10% or 20% increase in daily number of contacts (Figure 2C, 2F); although the outbreak would last longer, compared to the scenario in which there was no change in contacts post-lockdown (Figure 1A). On day 250 (day 176 after the start of the vaccination campaign), vaccination coverage with only one dose was 48%, with coverage of 21% and 20% for both doses of Pfizer-BioNTech and Moderna vaccines, respectively. These results highlight the importance of maintaining NPIs to allow for a sufficient increase in population-level immunity through vaccination prior to lifting public health measures.

## Discussion

Our results indicate that vaccination could have a large impact on reducing hospitalizations and deaths in Ontario, Canada. However, achieving this impact is contingent upon continued population-wide NPIs during the immunization program until there is adequate vaccination coverage. We found that without sufficient population immunity, even a 10% increase in daily contacts would reverse the declining trend of the outbreak achieved during the lockdown. Our results further highlight the importance of an accelerated vaccination rollout to prevent an outbreak surge that may arise due to the erosion of adherence to NPIs.

With the current and expected increases in daily vaccination rates, our results show that lifting restrictive measures before June 2021 is likely to trigger a resurgence of infections. This timeline is particularly important when considering the presence of new SARS-CoV-2 variants that have shown significantly higher transmissibility and risk of deaths compared to the original pandemic strain (41–44). Analysis of samples from Greater Toronto Area, the largest metropolitan population in Ontario, show an increase of over 6-fold in the spread of variants of concern from mid-December 2020 to early February 2021 (45). The transmissibility of these variants relative to the original strain was estimated to exceed 1.4 (45). Although our model does not explicitly include these variants, increasing daily contact rates could be considered as a proxy for a higher virus transmissibility. With over 30% positivity rate for variants of concern as of March 13, 2021 in Ontario (46), early relaxation of restrictive measures could lead to uncontrollable outbreaks as a result of reduced social distancing and wider spread of these variants.

Our results should be interpreted within the context of study assumptions and limitations. First, we assumed that vaccines would continue to be distributed according to the tested schedules in clinical trials. Recent evidence concerning vaccine efficacy after a single dose and with a delayed second dose (47–49) have led to changes in recommendations by Canada’s National Advisory Committee on Immunization, extending dose intervals for COVID-19 vaccines to optimize early vaccine rollout. A delay in administering the second dose of vaccines would affect the overall level of population immunity and, therefore, warrants further investigation on the effect of timing and the degree to which NPIs are lifted on outbreak control. Second, we did not consider drop-out over the course of the vaccination program, which would affect the level of protection conferred by vaccination. Third, the daily rate of vaccination was parameterized based on administered doses thus far and the expected increase in vaccine distribution in the coming months. Fourth, the inclusion of Oxford-AstraZeneca and Janssen vaccines in rollout could affect our results due to differences in vaccine efficacies, number of doses, and uptake of these vaccines. Despite the proven safety and efficacy of current vaccines, vaccine hesitancy (50) may result in a slower increase in the vaccination coverage than assumed in our model.

Therefore, a highly efficient national adverse event reporting system will be critical to supporting public confidence and improving vaccine uptake.

Notwithstanding these considerations, our study indicates that, while vaccination with highly effective vaccines could significantly mitigate COVID-19 outbreaks, it is unlikely to eliminate the need for NPIs before a sufficiently high level of population immunity is achieved. Our results show that vaccination is a key public health measure in the fight against the COVID-19 pandemic.

## Data Availability

The model was coded in Julia language and is available at: https://github.com/thomasvilches/vac_covid_ontario.

