## Supplementary File for "Projecting the impact of a two-dose COVID-19 vaccination campaign in Ontario, Canada"

This appendix provides further details of model structure and parameterization, and estimates for the reduction of overall attack rate and disease outcomes with the accelerated vaccination campaign.

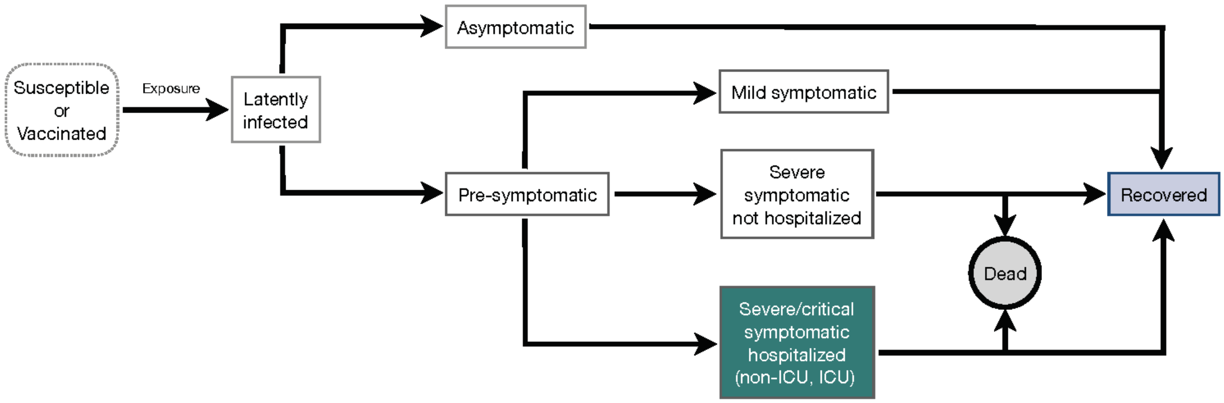


**Figure A1.** Schematic model diagram for infection and natural history of disease.

**Table A1.** Percentage of population in different age groups with and without comorbidities.

| Condition | 0-4 | 5-17 | 18-39 | 40-59 | 60-79 | 80+ | All age groups |
|---|---|---|---|---|---|---|---|
| With comorbidity | 5.0% | 10% | 18% | 38% | 60% | 72% | 32% |
| Without comorbidity | 95% | 90% | 82% | 62% | 40% | 18% | 68% |
| **Fraction of total population** | 4.8% | 15% | 29% | 27% | 20% | 4.2% | 100% |

**Table A2.** Mixing patterns and the daily number of contacts derived from empirical observations. Daily numbers of contacts were sampled from negative binomial distributions for different scenarios.

| Age group | Proportion of contacts between age groups | | | | | Daily number of contacts with stay-at-home order Mean (SD) | Daily number of contacts for self-isolated individual Mean (SD) |
|---|---|---|---|---|---|---|---|
| | 0-4 | 5-19 | 20-49 | 50-65 | 65+ | | |
| **0-4** | 0.2287 | 0.1839 | 0.4219 | 0.1116 | 0.0539 | 2.86 (2.14) | 0.8 (2.14) |
| **5-19** | 0.0276 | 0.5964 | 0.2878 | 0.0591 | 0.0291 | 4.70 (3.28) | 1.32 (3.28) |
| **20-49** | 0.0376 | 0.1454 | 0.6253 | 0.1423 | 0.0494 | 3.86 (2.95) | 1.08 (2.95) |
| **50-65** | 0.0242 | 0.1094 | 0.4867 | 0.2723 | 0.1074 | 3.15 (2.66) | 0.88 (2.66) |
| **65+** | 0.0207 | 0.1083 | 0.4071 | 0.2193 | 0.2446 | 2.24 (1.95) | 0.63 (1.95) |

**Table A3.** Vaccination coverage of different age groups and population segments with two doses of vaccines.

| | Age group | | | Individuals with comorbidities | Individuals without comorbidities | Healthcare workers |
|---|---|---|---|---|---|---|
| | 18-19 | 20-64 | 65+ | | | |
| Vaccine coverage | 37% | 47% | 70% | 56% | 34% | 90% |

**Table A4.** Estimated vaccine efficacies with associated timelines.

| Vaccine efficacy | Weeks after the first dose | | Weeks after the second dose | |
|---|---|---|---|---|
| Pfizer-BioNTech | 1-2 | 3 | 1 | >1 |
| Infection | None | 46% (40% - 51%) | 60% (53% - 66%) | 92% (88% - 95%) |
| Symptomatic disease | None | 57% (50% - 63%) | 66% (57% - 73%) | 94% (87% - 98%) |
| Severe disease | None | 62% (39% - 80%) | 80% (59% - 94%) | 92% (75% - 100%) |
| Moderna | 1-2 | 3-4 | 1-2 | >2 |
| Infection | None | 61% (31% – 79%) | 61% (31% – 79%) | 93.5% (85.2% - 97.2%) |
| Symptomatic disease | None | 92.1% (68.8% - 99.1%) | 92.1% (68.8% - 99.1%) | 94.1% (89.3% - 96.8%) |
| Severe disease | None | 92.1% (68.8% - 99.1%) | 92.1% (68.8% - 99.1%) | 100% |

**Table A5.** Risk of death due to COVID-19.

| | Age groups | | | | | | |
|---|---|---|---|---|---|---|---|
| **Hospitalized cases** | **0-19** | **20-44** | **45-54** | **55-64** | **65-74** | **75-84** | **85-100** |
| Non-ICU | 0.1% | 0.15% | 0.65% | 1.0% | 2.0% | 7.35% | 38.0% |
| ICU | 0.2% | 0.22% | 0.8% | 2.2% | 4.0% | 8.0% | 40.0% |

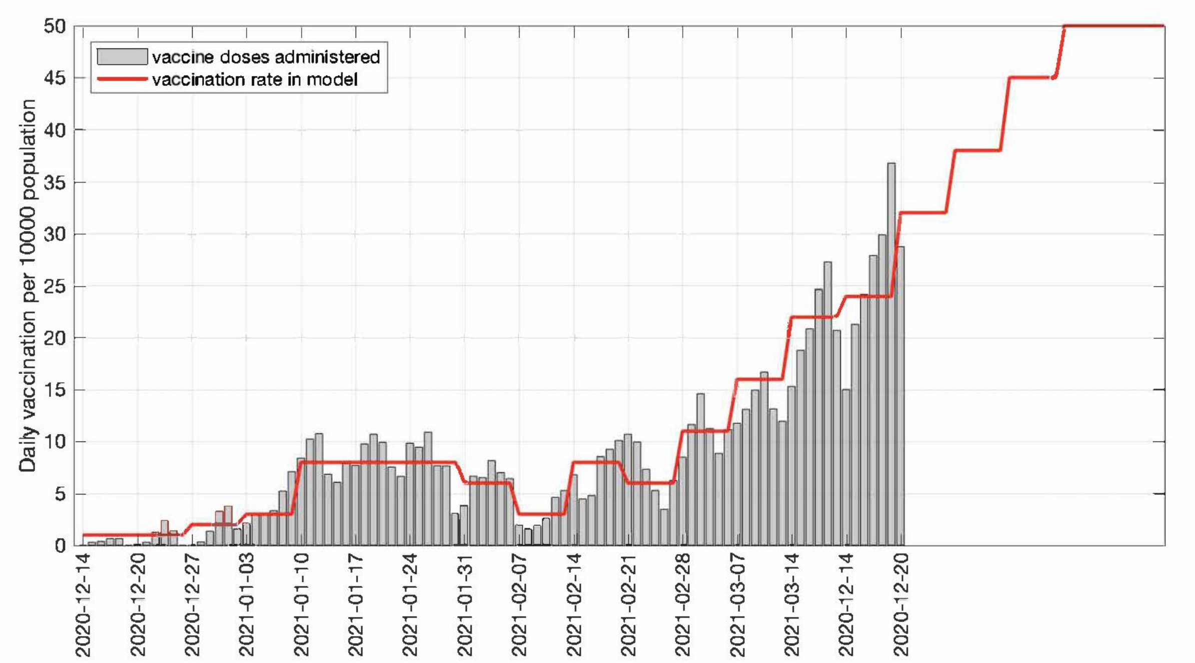


**Figure A2.** Daily vaccination rate.
